# Depression events associated with proton-pump inhibitors in postmarketing drug surveillance data

**DOI:** 10.1101/2023.06.03.23290931

**Authors:** Tigran Makunts, Haroutyun Joulfayan, Kenneth Ta, Ruben Abagyan

**Author notes:** Correspondence to RA.

## Abstract

Proton-pump inhibitors, PPIs, are widely prescribed and are available over the counter for prolonged reduction of stomach acid production and related disorders. PPIs irreversibly inhibit the hydrogen/potassium ATPase in gastric parietal cells. Recent retrospective studies have described an association between PPI use and depression. However, there is conflicting evidence that PPI therapy improves depressive symptoms. Considering the widespread use and over the counter availability of these drugs, further investigation into depression adverse event was warranted with a larger scale postmarketing set of reports. Here we analyzed over 68,178 reports from the FDA Adverse Event Reporting System consisting of PPI and histamine-2 receptor antagonist monotherapy records and found a statistically significant association between use of PPIs and depression. Additionally, we analyzed each of the six currently marketed PPIs individually and observed the association with the depression adverse reaction for all of them.

## Introduction

Proton-pump inhibitors (PPIs) are commonly used to treat acid related disorders such as Helicobacter Pylori induced gastric ulcers, gastroesophageal reflux disease (GERD), erosive esophagitis, and Zollinger-Ellison syndrome^1-3^. PPIs have therapeutic superiority to histamine-2 receptor antagonists (H2RAs)^4^ due to their irreversible inhibition of the H+/K+ ATPase in gastric parietal cells^5,6^. However, PPI pharmacology may not be limited to localized parietal cell action^7,8^. Despite efforts to minimize over-prescription of these medications, PPI use has been steadily increasing^9^. According to National Health and Nutrition Examination Survey, the number of PPI prescriptions among 40–64 year old individuals had increased from 1999 to 2012^10^, even though these studies did not account for over-the-counter (OTC) use of PPIs.

Currently, there are six drugs of the PPI class approved by the U.S. food and drug administration: rabeprazole, pantoprazole, dexlansoprazole, lansoprazole, esomeprazole, and omeprazole. The latter three are available OTC without any restrictions.

Common adverse drug reactions (ADRs) associated with PPIs observed in controlled trials include nausea, diarrhea, flatulence, vomiting and headache. Serious ADRs include throat tightness, rash, and difficulty breathing^11-16^. Post-approval studies have found additional association of PPI use with the following ADRs: *Clostridium difficile* associated diarrhea, bone fractures, and hypomagnesemia, and are now listed as precautions in the FDA-package inserts. Additionally, recent studies analyzing the FDA Adverse Event Reporting System (FAERS) data have observed multiple electrolyte abnormalities and a broad spectrum of neurological disorders disproportionally reported after PPI use when compared to H2RAs ^17,18^.

The physiology behind neurological and psychiatric conditions is often shares common molecular mechanisms^19,20^, resulting in depression’s comorbidity with neurological disorders^21^. In an epidemiological study by Laudisio et al., PPI use was associated with depression in a geriatric population, while the association was not observed in the H2RA cohort^22^. In another study conducted in Sweden by Wang et al.,^23^ PPI use was associated with increased risk of anxiety and depression in children. Furthermore, a Taiwanese nationwide population-based study by Huang et al.^24^ confirmed this association. There is conflicting evidence suggesting improvement of depressive symptoms following PPI therapy^25^.

Depression was declared by the World Health Organization (WHO) to be the 3rd greatest cause of burden for disease worldwide^26^, and considering the common use of PPIs, this association needed to be further investigated. In this study we performed analysis of millions of the FAERS reports and identified a significant increase in depression reports in patients taking PPIs as monotherapy when compared to H2RAs related reports.

## Methods

### FDA adverse event reporting system

The FDA Adverse Event Reporting System (FAERS) and its older version AERS contain the FDA’s post-marketing drug and biologic product safety data. Reports are submitted by product vendors, and, on a voluntary basis, by legal representatives, healthcare professionals and consumers to the FDA through MedWatch^27,28^, the FDA Safety Information and Adverse Event Reporting Program. Pharmaceutical industry/manufacturers are legally required to forward the information to the FDA.

Over 17.8 million FAERS/AERS reports, collected from January 2004 to July 2022, were used for the analysis. Data sets are available online at: https://www.fda.gov/Drugs/GuidanceComplianceRegulatoryInformation/Surveillance/AdverseDrugEffects/ucm082193.htm

### Data preparation

Quarterly FAERS/AERS ASCII reports were downloaded in their original format. The data form different years were unified into a consistent format. Additionally, since the ADR reports were collected from around the world, it was necessary to translate drug brand names into generic ones using online drug databases. A total of 17,828,284 combined FAERS/AERS reports were used for the analysis^29,30^. The reports were further narrowed down to exclude reports submitted by legal representatives, consumers, and legal representatives to avoid potential bias.

### Analysis and control cohort selection

FAERS/AERS Reports where PPIs and H2RAs were used as monotherapy were selected into the respective cohorts. The term “monotherapy” pertains to records where only a single treatment is listed in the report. PPIs-monotherapy cohorts collectively included 7,790 reports, and H2RA-monotherapy cohorts included 60,388 reports.

Reporting odds ratio (ROR) analysis was performed by comparing the reported PPIs ADRs in relation to H2RAs report numbers with and without an ADR of interest. The PPI monotherapy cohort was further split into individual PPI cohorts which included omeprazole (n□=□2,429) esomeprazole (n□=□1,843), pantoprazole (n□=□1,674), lansoprazole (n□=□1,063), Dexlansoprazole (n=378), and rabeprazole (n□=□403). Each individual PPIs reported depression ADRs were calculated and compared to the H2RA cohort to screen for potential ADR variability within individual PPIs in the cohort. Demographic analysis was performed (See Supplement).

### Statistical analysis

#### Descriptive statistics

Frequency for depression ADR was calculated by the equation:

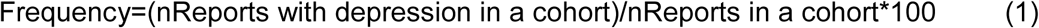

#### Comparative statistics

Depression report rates were compared via the Reporting Odds Ratio (ROR) analysis:

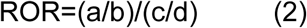

Where,

Number of cases in PPI group with depression

Number of cases in PPI group with no depression

Number of cases in H2RA group with depression

Number of cases in H2RA group with no depression

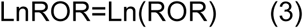

Standard Error of Log Reporting Odds Ratio;

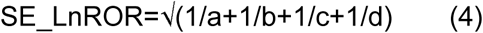

95% Confidence Interval;

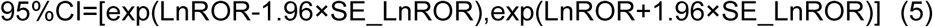

Haldane-Anscombe correction was not applied to cohorts with zero depression reports ^31^.

## Results

Combined PPIs monotherapy reports had a significantly higher number of depression ADRs in comparison with H2RAs monotherapy ones, with ROR being 40.57, 95% CI [20.61, 79.87]). When studied individually, five out of six PPI monotherapy cohorts had a significant increase in the number of reported depression ADRs in comparison with the H2RA control cohort. The ROR values were as follows: omeprazole (ROR 60.25, 95% CI [28.78, 126.14]), pantoprazole (ROR 39.94, 95% CI [16.94, 94.17]), rabeprazole (ROR 91.25, 95% CI [33.01, 252.28]), esomeprazole (ROR 29.63, 95% CI [12.03, 73.00]), and dexlansoprazole (ROR 16.02, 95% CI [2.04, 125.43]). The lansoprazole cohort did not meet the 95% CI significance criterion for reported depression risk (ROR 5.69, 95% CI [0.73, 44.45]) (Figs. 1 and 2).

**Figure 1.**
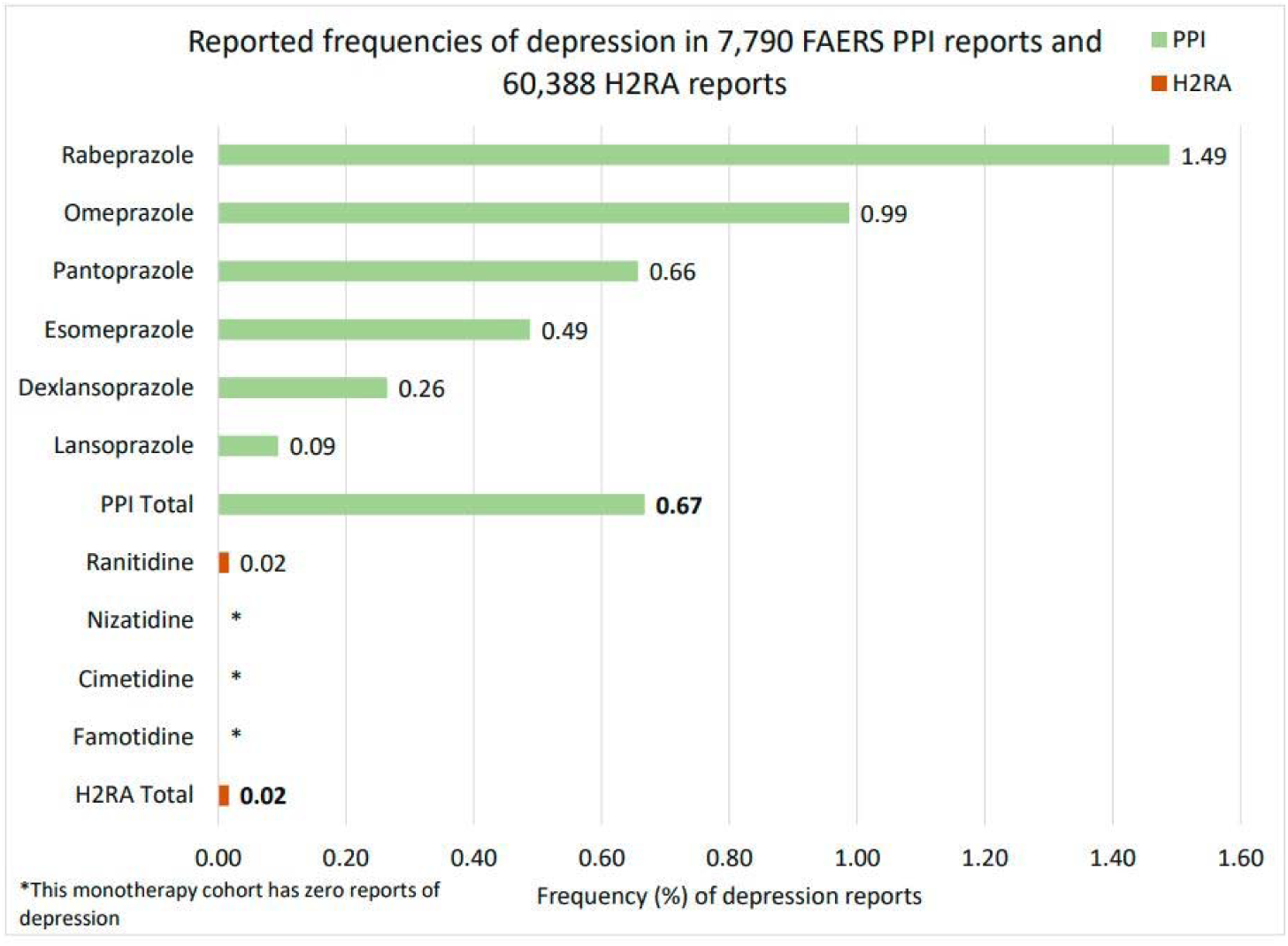
Reported frequencies of depression ADRs for patients in FAERS/AERS who took PPIs (n=7,790) and H2RAs (60,388) as monotherapy.

**Figure 2.**
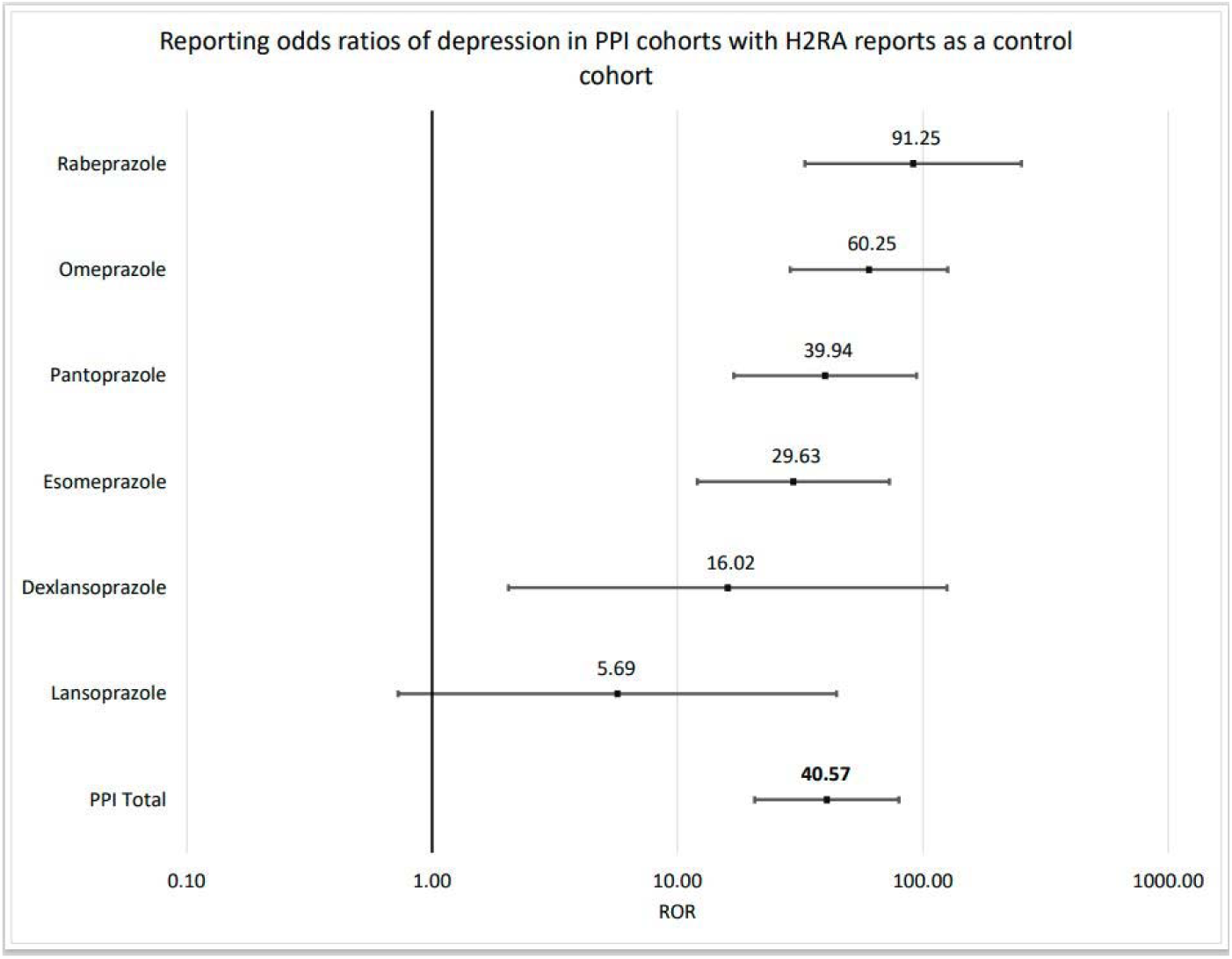
Reporting odds ratios (ROR) of for patients in FAERS/AERS who took PPIs (n=7,790) and H2RAs (60,388) as monotherapy. PPI monotherapy cohort was further split into individual PPI cohorts which included omeprazole (n□=□2,429) esomeprazole (n□=□1,843), pantoprazole (n□=□1,674), lansoprazole (n□=□1,063), dexlansoprazole (n=378), and rabeprazole (n□=□403) for individual ROR analysis. X-axis is presented in log scale.

### Depression reports in males

There was a significant increase in the number of reported depression ADRs in the combined PPI monotherapy cohort when compared to H2RAs monotherapy cohort (OR 39.38, 95% CI [12.95, 119.71]) when only male patients taking PPIs were analyzed.

Each of the individual PPI treatment analyzed had a significant increase in the number of depression ADRs in comparison with H2RAs control, with the exception of lansoprazole and dexlansoprazole which had zero depression ADR reports. The ROR values were as follows: omeprazole (ROR 30.58, 95% CI [7.64, 122.46]), pantoprazole (ROR 84.27, 95% CI [23.72, 299.41]), rabeprazole (ROR 53.77, 95% CI [5.98, 483.93]), esomeprazole (ROR 35.95, 95% CI [8.03, 160.96] (Figs. 3 and 4).

**Figure 3.**
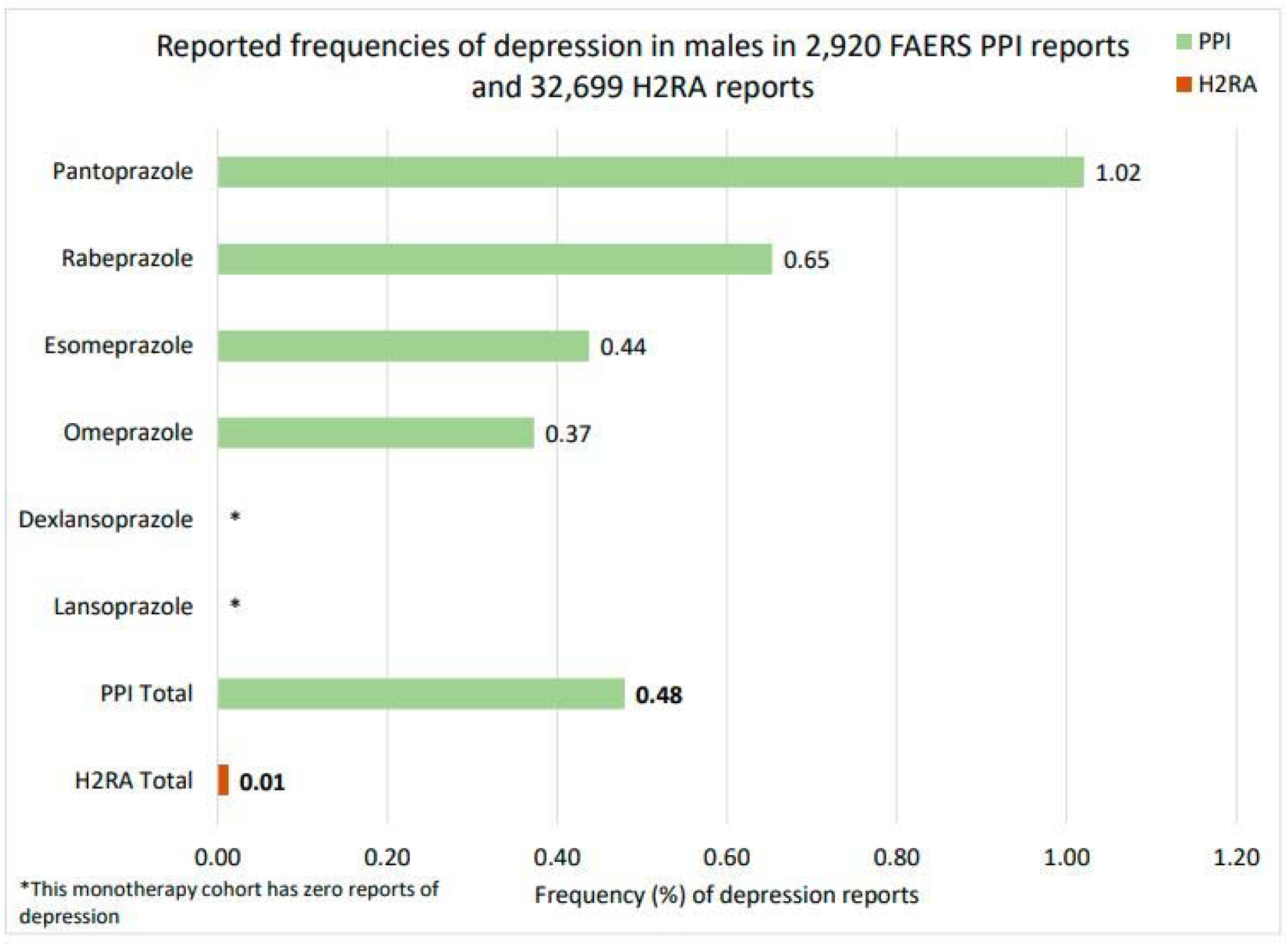
Reported frequencies of depression ADRs for male patients in FAERS/AERS who took PPIs (n=2,920) and H2RAs (32,699) as monotherapy.

**Figure 4.**
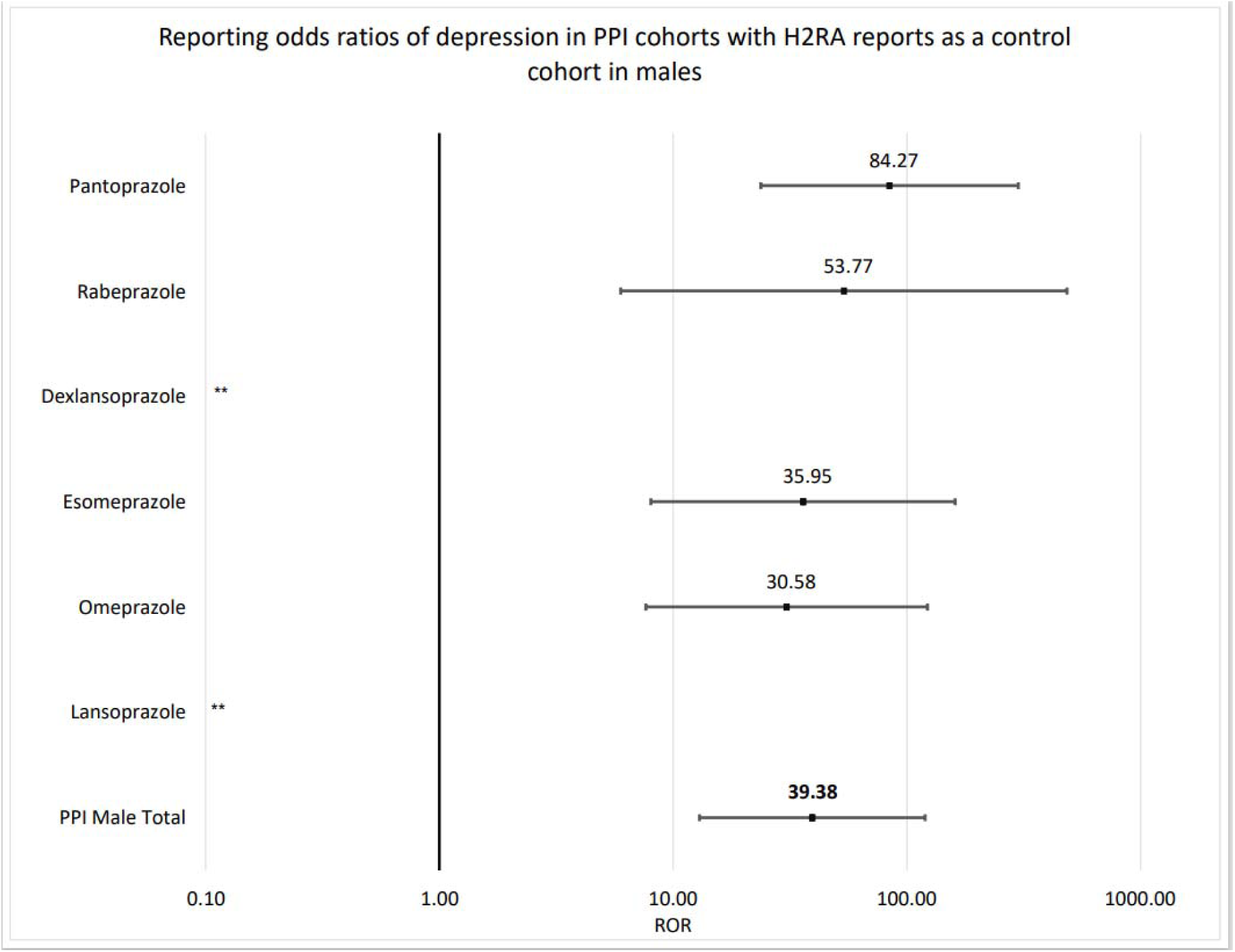
Reporting odds ratios (ROR) of for male patients in FAERS/AERS who took PPIs (n=2,920) and H2RAs (32,699) as monotherapy. PPI monotherapy male cohort was further split into individual PPI cohorts which included omeprazole (n□=□1,073) esomeprazole (n□=□685), pantoprazole (n□=□588), lansoprazole (n□=□322), dexlansoprazole (n=99), and rabeprazole (n□=□153) for individual ROR analysis. X-axis is presented in log scale. **Lansoprazole and dexlansoprazole cohorts had no depression ADR reports.

**Figure 5.**
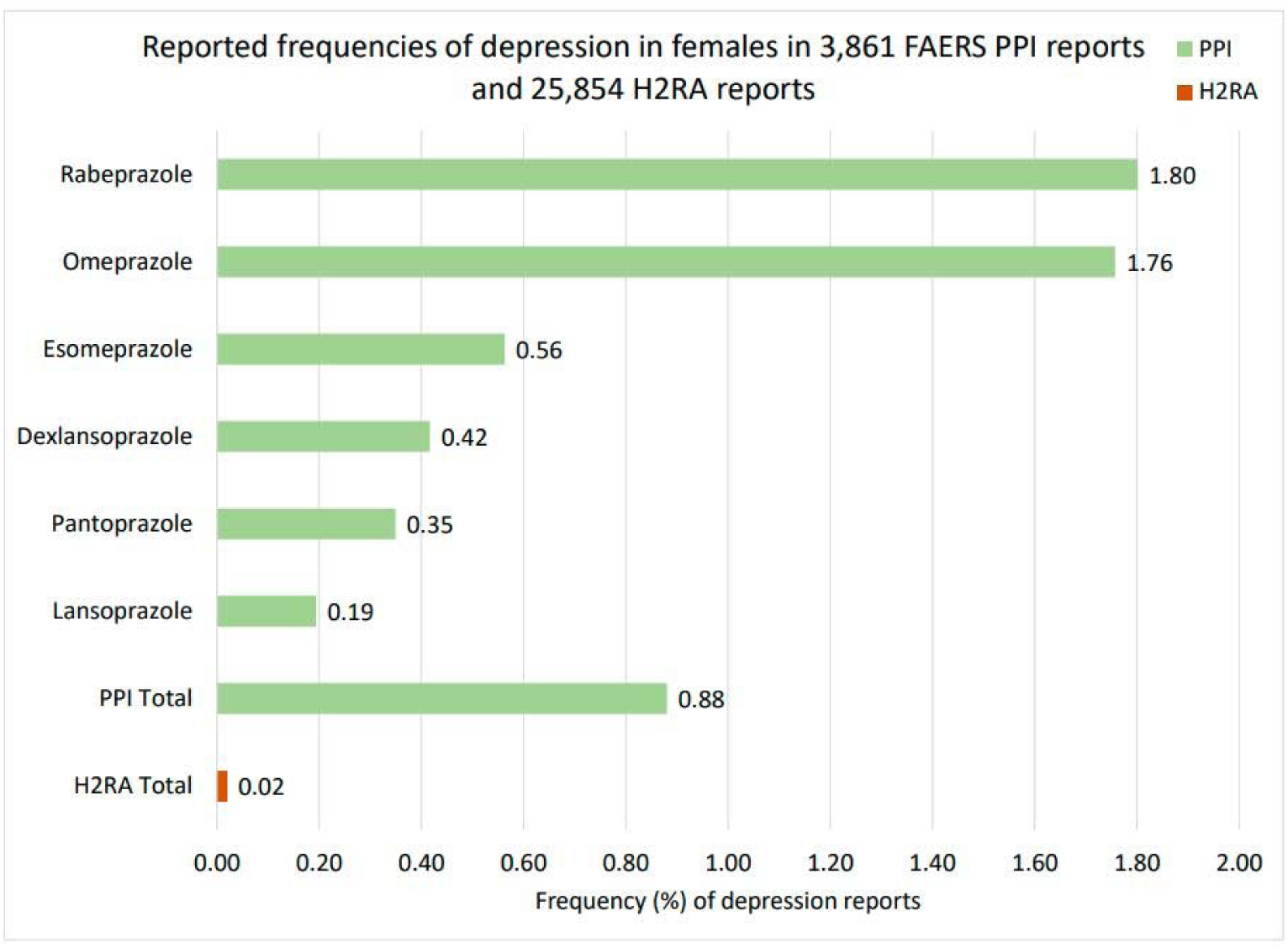
Reported frequencies of depression ADRs for female patients in FAERS/AERS who took PPIs (n=3,861) and H2RAs (25,854) as monotherapy.

**Figure 6.**
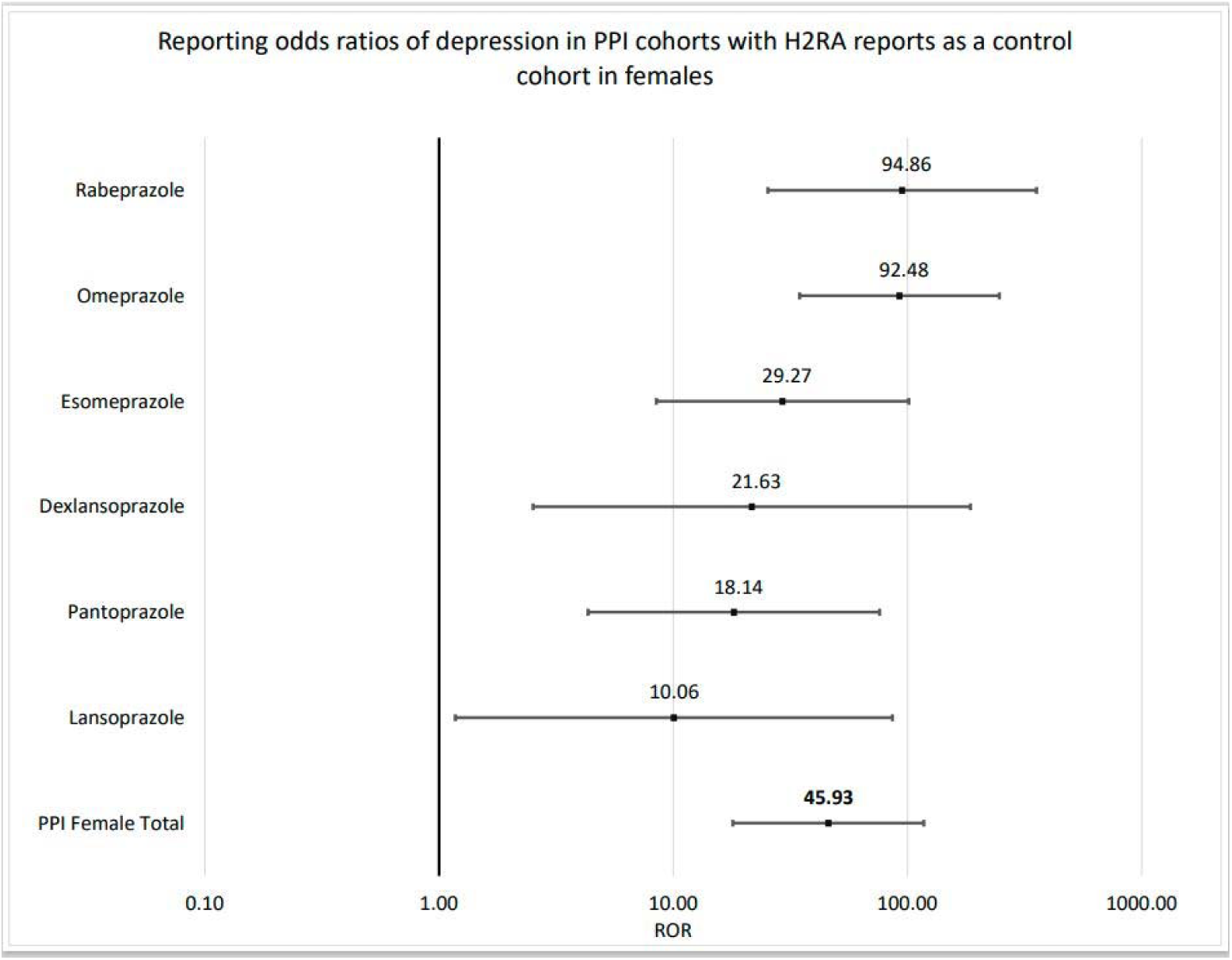
Reporting odds ratios (ROR) of for female patients in FAERS/AERS who took PPIs (n=3,861) and H2RAs (n=25,854) as monotherapy. PPI monotherapy female cohort was further split into individual PPI cohorts which included omeprazole (n=1,138) esomeprazole (n=888), pantoprazole (n=858), lansoprazole (n=515), dexlansoprazole (n=240), and rabeprazole (n=222) for individual ROR analysis. X-axis is presented in log scale.

### Depression reports in females

There was a significantly higher number of depression ADR reports in the combined PPI monotherapy female-only cohort when compared to H2Ras monotherapy cohort (OR 45.93, 95% CI [17.95, 117.51]). All individual PPIs cohorts had a significant increase in the number of depression ADR reports when compared to H2RA control: omeprazole (ROR 92.48, 95% CI [34,65, 246.87]), pantoprazole (ROR 18.14, 95% CI [4.32, 76.03]), rabeprazole (ROR 94.86, 95% CI [25.30, 355.65]), esomeprazole (ROR 29.27, 95% CI [8.46, 101.30]), dexlansoprazole (ROR 21.63, 95% CI [2.52, 185.86]), and lansoprazole (ROR 10.06, 95% CI [1.17, 86.25]).

## Discussion

In this study, we quantified and confirmed a statistically significant association between PPI exposure and depression ADRs using 68,178 PPI and H2RA ADR reports from the FAERS/AERS database. To our knowledge, it is the first study investigating PPI relationship to depression utilizing the most recent population-scale postmarketing surveillance data.

Although the number of depression reports was relatively low, ranging from 0.1% to 1.49% of PPI monotherapy ADR reports. The depression ADR ROR numbers were statistically significant when compared to the H2RA control cohort. This study expanded on the previous epidemiological evidence provided in the geriatric study by Laudisio et al.^22^ and the pediatric study by Wang et al^23^. Additionally, in our study depression ADRs were analyzed across age groups. The depression ADR signal prevailed in all groups above 20-29 to >= 70 year old cohorts (Supplement S2).

The molecular mechanisms of the observed association may originate from several pathways and are beyond the scope of this study. Due to complex pharmacology of PPIs it is difficult to pinpoint a single physiological mechanism involved in the etiology of depression ADR. In a previously published mini review we discussed some of the mechanisms by which PPIs might be causing neurological adverse events^32^, including hypomagnesemia and impaired vitamin B absorption.

A systematic review published by Derom et al.^33^ found an association between higher dietary magnesium intake and lower depression symptoms. In another publication by Cheungpasitporn et al.^34^ the investigators evaluated the effect of high magnesium and found a similar association. Magnesium is known to play multiple important roles in the central nervous system (CNS)^35^, and inadequate levels affecting psychiatric symptoms seem intuitive.

In a four-year longitudinal study by Laird et al.^36^ in adults 50 years old or higher, vitamin B12 deficiency was found to be associated with increased depressive symptoms. In another study by Syed et al.^37^, when supplemented along with antidepressants, vitamin B12 significantly decreased depressive symptoms. A systematic review by Almeida et al.^38^ has found that the severity of depression symptoms in not decreased over a short period. However, there are long term benefits in vitamin B12 supplementation for depression management.

Coincidentally, the PPI users reported both hypomagnesemia and vitamin B12 deficiency. The report counts were as follows: 35 reports of decreased vs. 1 report of increased magnesium levels, and 43 reports of decreased vs 1 report of increased vitamin B12 levels (Supplement S2 and S3). This associations of PPIs with vitamin B12 and magnesium absorption abnormalities^39,40^ are plausible mechanisms that could explain the elevated reporting numbers of depression ADRs. Moreover, regardless of specific molecular mechanisms related to depression ADR, it is not surprising to see psychiatric ADRs, given the previous evidence of a wide range of neurological damage associated with the PPI use^18,41-44^.

The observed increased risk of depression with use of PPIs calls for more careful consideration in using PPIs for people at high risk for depression. PPIs should be used for the shortest time necessary as recommended by the FDA and other regulatory authorities.

### Study Limitations

Only a fraction of population incidences is represented in FAERS/AERS due to the mostly voluntary nature of the reporting. FAERS/AERS reporting can be biased based on multiple factors such as legal challenges or news events influencing submissions^45^. Additionally, significant underreporting AEs has been observed depending of drug, type of an ADR and seriousness^46^. Causality of depression cannot be inferred from the association found in this study since the cases were not individually adjudicated by a clinician. However, the study provides a statistically significant signal that needs to be further investigated in a controlled setting.

## Data Availability

All data produced in the present work are contained in the manuscript.

## Data availability

The data sets available online to the public are de-identified. Institutional Review Board requirements do not apply under 45 CFR 46.102. https://www.fda.gov/drugs/questions-and-answers-fdas-adverse-event-reporting-system-faers/fda-adverse-event-reporting-system-faers-latest-quarterly-data-files. Both FAERS and AERS datasets are de-identified and are made available online at: http://www.fda.gov/Drugs/GuidanceComplianceRegulatoryInformation/Surveillance/AdverseDrugEffects/ucm082193.htm. Institutional Review Board Requirements do not apply under 45 CFR 46.102. There was no direct human participation in the study. All experiments were performed in accordance with relevant guidelines and regulations.

## Acknowledgements

We thank Drs. Rabia Atayee and Isaac V. Cohen for helpful discussions.

## Author information

### Contributions

H.J. performed the experiments, R.A. and T.M. designed the study and, K.T., H.J., R.A., and T.M. drafted the manuscript and reviewed the final version. R.A. processed the data set.

