## Supplement for "Depression events associated with proton-pump inhibitors in postmarketing drug surveillance data"

**Supplement S1. Demographic analysis**

Sex

|  | PPI monotherapy | H2RA monotherapy | PPI depression | H2RA depression |
| --- | --- | --- | --- | --- |
| Total reports | 100% (7790) | 100% (60388) | 0.74% (58) | 0.02% (11) |
| Male | 37.48% (2920) | 54.15% (32699) | 0.48% (14) | 0.02% (4) |
| Female | 49.56% (3861) | 42.81% (25854) | 0.88% (34) | 0.02% (5) |
| Unspecified sex | 12.95% (1009) | 3.04% (1835) | 0.4% (4) | 0.05% (1) |
| χ-squared male vs. female | 2.94 | 3.00 | 0.18 | 0 |

Age

|  | PPI monotherapy | H2RA monotherapy | PPI depression | H2RA depression |
| --- | --- | --- | --- | --- |
| Total reports | 100% (7790) | 100% (60388) | 0.67% (52) | 0.02% (10) |
| Invalid ages (<1 y.o.) | 1.45% (113) | 0.15% (88) | 0% (0) | 0% (0) |
| Unspecified ages (empty) | 33.11% (2579) | 1.41% (849) | 0.27% (7) | 0.12% (1) |
| 1-9 y.o. | 1.42% (111) | 0.1% (58) | 0% (0) | 0% (0) |
| 10-19 y.o. | 1.91% (149) | 0.2% (121) | 0% (0) | 0.83% (1) |
| 20-29 y.o. | 3.22% (251) | 1.36% (819) | 0.8% (2) | 0% (0) |
| 30-39 y.o. | 5.94% (463) | 5.55% (3350) | 1.3% (4) | 0.03% (1) |
| 40-49 y.o. | 8.2% (639) | 16.49% (9956) | 1.1% (7) | 0.01% (1) |
| 50-59 y.o. | 11.96% (932) | 32.49% (19621) | 1.72% (16) | 0.02% (4) |
| 60-69 y.o. | 12.79% (996) | 29.41% (17758) | 1% (10) | 0% (0) |
| >= 70 y.o. | 19.99% (1557) | 12.86% (7768) | 0.39% (6) | 0.03% (2) |

Country

|  | PPI monotherapy | H2RA monotherapy | PPI depression | H2RA depression |
| --- | --- | --- | --- | --- |
| Total reports | 100% (7790) | 100% (60388) | 0.67% (52) | 0.02% (10) |
| United States | 50.96% (3970) | 98.83% (59681) | 0.65% (26) | 0.02% (10) |
| France | 8.54% (665) | 0.03% (18) | 1.65% (11) | 0% (0) |
| United Kingdom* | 6.98% (544) | 0.11% (68) | 1.29% (7) | 0% (0) |
| Japan | 5.66% (441) | 0.47% (284) | 0% (0) | 0% (0) |
| Italy | 4.35% (339) | 0.11% (66) | 0.29% (1) | 0% (0) |
| Canada | 3.2% (249) | 0.03% (19) | 0.8% (2) | 0% (0) |
| Germany | 2.84% (221) | 0.03% (20) | 0.45% (1) | 0% (0) |
| Spain | 2.41% (188) | 0.04% (23) | 0% (0) | 0% (0) |
| Turkey | 1.78% (139) | 0.02% (10) | 0.72% (1) | 0% (0) |
| Brazil | 1.68% (131) | 0% (2) | 1.53% (2) | 0% (0) |
| Other countries | 11.59% (903) | 0.33% (197) | 0.11% (1) | 0% (0) |

*Combined the gb and uk codes into United Kingdom

**Supplement S2. B12 abnormality reports**

| Adverse event | n(%) |
| --- | --- |
| Vitamin B12 increased | 1(0.01) |
| Vitamin B12 abnormal | 2(0.03) |
| Anaemia vitamin B12 deficiency | 5(0.06) |
| Vitamin B12 decreased | 8(0.10) |
| Vitamin B12 deficiency | 28(0.36) |

**Supplement S3. Magnesium abnormality reports**

| Adverse event | n(%) |
| --- | --- |
| Blood magnesium increased | 1(0.01) |
| Magnesium deficiency | 2(0.03) |
| Blood magnesium decreased | 33(0.42) |
